# Correlates of long-acting reversible contraceptive (LARC) use among young women in Southern Africa: a secondary analysis from HPTN 082

**DOI:** 10.1101/2025.09.16.25335943

**Authors:** Phionah Kibalama Ssemambo, Michael Burton, Brenda Gati Mirembe, Clemensia Nakabiito, Deborah Donnell, Geetha Beauchamp, Sinead Delany-Moretlwe, Connie Celum, Jennifer Velloza

## Abstract

**Background:** Long-acting reversible contraception (LARCs), including intrauterine devices (IUDs), injectables, and implants, are highly effective in preventing unintended pregnancies but LARC use rates are low among African adolescents and young women (AGYW). Understanding factors associated with LARC uptake and continuation among African AGYW may provide insights into strategies to promote LARC use.

**Methods:** We conducted a secondary data analysis from the HIV Prevention Trials Network (HPTN 082) pre-exposure prophylaxis (PrEP) study, which enrolled 451 AGYW in Zimbabwe and South Africa ages 16-25 years, who reported vaginal or anal sex in the prior month, and reported PrEP interest (ClinicalTrials.gov, NCT02732730). Contraception and contraceptive counseling were offered at enrollment and visits at 4, 8, 13, 26, and 39 weeks post enrollment, with follow-up through 52 weeks. The outcome variable was any LARC use, defined as copper or hormonal IUDs, injectable contraceptives, and implants. We performed descriptive analyses and regression models to assess contraceptive use patterns and characteristics associated with LARC use and condomless sex.

**Results:** Overall, 60% (299/499) of AGYW adopted a LARC method at enrollment and 78% (234/299) persisted on a LARC method during follow up. Among the 449 women who used contraception at enrollment and/or during follow-up, 38 (8.4%) switched between non-LARC to LARC and 34 (7.5%) discontinued contraception at some point during the study. AGYW not using a LARC at enrollment were more likely to switch contraceptive method through week 39 compared to those already using a LARC (32.7% vs. 14.7%, respectively; p-value<0.001). Factors significantly associated with choosing a LARC method were prior pregnancy [adjusted odds ratio [aOR]:2.46; 95% confidence interval [CI]: 1.59-3.79; p<0.01], and comfort talking to close friends about sexual relationships [aOR:1.82; 95% CI:1.02-3.23; p=0.04]. Consistent condom users were less likely to choose a LARC method [aOR:0.27; 95% CI:0.19-0.39; p<0.01].

**Conclusion:** Contraceptive counseling and offering LARC methods with HIV PrEP was associated with a majority of African AGYW selecting a LARC method. Peer support is important in facilitating LARC use and the high contraceptive efficacy of LARC should be discussed with AGYW using condoms for contraception. Contraceptive counseling and promotion of LARCs should be integrated with PrEP delivery for African AGYW.

## Introduction

Unintended pregnancy remains an alarming global public health problem. By 2019, WHO estimated 10 million unintended pregnancies annually among AGYW aged 15-24 years in sub-Saharan Africa (SSA) [1-3]. Given the continued growth of the adolescent population globally, adolescent pregnancies are also projected to increase, with SSA experiencing the most substantial increases [4-6]. Early and unintended pregnancies among adolescents are linked to serious medical complications, including eclampsia, endometritis, unsafe abortions, maternal death, and disabilities such as obstetric fistula. These pregnancies also lead to adverse infant outcomes [5-7] and significant socioeconomic consequences, such as disrupted education and diminished future opportunities for the young mothers [5, 8-10].

The use of effective contraceptive methods could significantly reduce unintended pregnancies among AGYW by up to 25% [11-13]. Effective contraceptive methods include short-acting (condoms, hormonal pills, patch, vaginal ring, and spermicides), permanent (tubal-ligation or vasectomy), and LARC which include hormonal implants, bimonthly or trimonthly injectables, and intrauterine devices (IUCDs) [14-18]. LARCs are more effective than shorter-term contraceptives, offering high user acceptability, low discontinuation rates, reversibility, and minimal side effects [19-21].

They require less partner involvement, ensure return to fertility upon removal, need no user adherence, and reduce health facility visits, thus easing the strain on the healthcare system [22-27]. LARCs, however, provide no protection against STIs including HIV. Condom use remains the most effective behavioural method for the prevention of HIV and unplanned pregnancies, requiring dual-method use of both LARC and condoms [28-30].

Despite the significant health and social benefits of LARC, its uptake among AGYW in SSA remains critically low, estimated at below 22% [31, 32]. This low uptake contributes to an unmet need for effective family planning, which stands at approximately 23% among AGYW in low- and middle-income countries [33]. Various factors influence contraceptive use among adolescents, including individual-level, societal, health system, socio-cultural, and religious influences such as peer pressure, insufficient parental counselling, low educational attainment, lack of comprehensive sexuality education, early sexual initiation, misconceptions about contraceptives, high costs, and non-friendly adolescent reproductive services [34]. There is a significant gap in understanding the specific barriers and facilitators affecting LARC uptake and persistence among African adolescents and young women, particularly within the context of regular contraceptive access, pre-exposure prophylaxis (PrEP) delivery, and sexually transmitted infection (STI) testing which offer opportunities to reach AGYW with routine sexual and reproductive health services. This research aims to address these gaps by exploring contraceptive method use and switching and facilitators of LARC use, among AGYW participating in an HIV PrEP demonstration project (“HPTN 082”) in southern Africa.

## Methods

### Parent Study Design

We conducted a secondary data analysis using data from the HPTN 082 trial to understand 1) prevalence and patterns of contraceptive use including initiations, discontinuations, and switches over time; 2) demographic and socio-behavioral characteristics associated with LARC use among AGYW; and 3) associations between LARC use and condomless vaginal sex to determine if LARC users need additional interventions to maintain protective sexual behaviors.

### Data source and access

The dataset for this secondary analysis was accessed for research purposes on 18 July 2023. Authors did not have access to information that could identify individual participants during or after data collection; all data were fully de-identified.

HPTN 082 was a Phase IV randomized prospective open-label PrEP demonstration trial conducted between 2016-2018, in South Africa (Cape Town and Johannesburg) and Zimbabwe (Harare) to assess PrEP acceptance and adherence among 451 HIV negative AGYW who were offered daily oral PrEP with adherence support [35]. Eligibility criteria for HPTN 082 included: not living with HIV, female at birth and aged 16-25 years, reported vaginal or anal sex in the month prior to screening, scored ≥ 5 in the VOICE risk behavior questionnaire [36], had regular access to phone with short message service (SMS) functionality, had normal renal function, was hepatitis B negative, and reported interest in taking PrEP. Pregnant women were excluded and women who became pregnant during follow-up discontinued PrEP.

PrEP was offered in conjunction with standard-of-care HIV prevention interventions (HIV testing, counseling, condoms, testing for STIs). Women had the option to initiate or decline PrEP at enrollment (decliners and any participants who initiated and then discontinued PrEP could initiate PrEP at any point until month 9).

Additional details on HPTN 082 primary objectives and study procedures are reported elsewhere [35]. Participants attended visits at weeks 4, 8, 13, 26, 39, and 52, during which HIV and pregnancy testing were conducted, and all women were followed for a total of 52 weeks. Contraceptive counseling was provided through week 39, allowing women the option to initiate, decline, switch, or discontinue a contraceptive method.

### Data Collection

The primary outcome variable was the use of LARCs, defined as the proportion of AGYW who were actively using LARCs such as intrauterine devices (IUDs), injectables, or contraceptive implants, during enrollment and follow-up visits. Contraceptive use data for a given visit was collected by asking participants about the contraception or family planning method(s) they were currently using, with the option to select multiple methods . Data on condomless vaginal sex was collected by asking about how often a condom was used during vaginal sex in the past month.

For this secondary analysis, we considered a range of sociodemographic and behavioral factors to be “exposures” given their potential influence on LARC use. At enrollment, demographic data on variables such as age and education were collected. Other socio-behavioral factors were assessed through self-reported measures at enrollment and then quarterly up to 52 weeks, via computer-assisted self-interviews (CASI). These factors included sexual behaviors, such as the number of sexual partners in the past 12 months and the age of sexual partners; condom use during vaginal or anal sex; history of intimate partner violence (IPV); social support from adults and close friends; comfort in discussing sexual relationships and health issues with friends; requests for assistance from friends; encouragement from close friends to start PrEP; hope for the future; perceived risk of pregnancy in the upcoming year; and history of past pregnancies. IPV was assessed using four items based on the World Health Organization’s IPV definitions. Participants who responded “yes” to at least one item were categorized as having experienced IPV. Social support was evaluated using two questions adapted from previous research with African AGYW, focusing on support from adults and close friends. Responses were scored on a Likert scale and summed to produce a total score ranging from 0 to 4, with higher scores indicating greater perceived social support. Hope for the future was assessed using six Likert scale items, with response options ranging from “I totally disagree,” “disagree,” “agree,” and “I totally agree,” to “prefer not to answer.” Participants’ scores ranged from 6 to 24, with higher scores indicating greater levels of hope [37]. Responses to data on condomless vaginal sex were categorized as Always, Often, Sometimes, Rarely, None, or Prefer not to answer. The explanatory variables were chosen based on their significant associations with contraceptive use identified in existing literature [11, 38-40].

### Data Analyses

The analysis included all women who were enrolled in HPTN 082 and initiated contraceptive use, while excluding the two participants who did not report contraceptive use during the follow-up period. Categorical variables were summarized using counts and percentages (N%) within each category, excluding the missing category from the denominator.

Descriptive statistics were first used to summarize sample characteristics, including LARC use and patterns of contraceptive switching at each study visit. For the analysis of switching behaviors, changes in contraceptive methods across study visits were represented using an alluvial plot. Participants were categorized into four distinct groups: 1) those not using contraceptives at enrollment but switched to a highly effective method, 2) those who started with a highly effective method but later discontinued, 3) those who switched between different highly effective methods, and 4) those who remained on the same highly effective method throughout the study. Highly effective contraception refers to methods that have a low failure rate when used correctly, typically defined as less than 1 pregnancy per 100 women per year. These methods include hormonal options like birth control pills, LARCs as well as permanent methods like sterilization. Contraceptive methods were condensed into categories, with participants assigned to their highest level of contraceptive use at each visit, ensuring they were counted only once per visit.

In this study, the term highest level refers to classifying contraceptive methods by effectiveness or type, assigning participants to the most effective method reported during a given visit. The paths in the plot, which illustrate transitions in contraceptive use over time, were color-coded based on the participant’s baseline contraceptive use. The width of each path was proportional to the number of participants, visually representing the flow and changes in contraceptive methods throughout the study.

Chi-square and Fisher’s exact tests were conducted to assess associations between baseline covariates and LARC use at baseline. For the primary analysis assessing associations between baseline covariates and LARC usage (yes/no) across visits up to week 39, we fit univariate and multivariate logistic regression models. Covariates with a two-sided p-value <0.10 in univariate models were included in the multivariate logistic regression models. Statistical significance in multivariate analyses was defined as a two-sided p-value <0.05, with an exchangeable correlation structure applied to all models.

To evaluate whether LARC use at enrollment was associated with condomless vaginal sex over the follow-up period, logistic regression models (univariate and multivariate multinomial) were fitted. Condomless vaginal sex was analyzed longitudinally through week 52 (including timepoints at Weeks 4, 8, 13, 26, 39, and 52), Adjusted odds ratios (aORs) with 95% confidence intervals (CIs) were calculated.

### Ethical considerations

The HPTN 082 study received Institutional Review Board approval from each study site. All methods were carried out in accordance with IRB approval, country, and local regulations. All participants provided written informed consent in English or in their preferred local language. Following local regulations, participants below the legal age for consent (age <18 years) provided assent, and parent or guardian informed consent was obtained.

## Results

Of the 449 AGYW included in the analysis, 55.5% (249/449) were aged 18–21 years, the median VOICE score was 7 (IQR: 6-8), the median number of sex partners in the previous 3 months was 1 (IQR: 1-2), half of the study participants reported an HIV-negative partner, and 70.1% reported that they did not engage in transactional sex. Two participants were excluded from the analysis as they did not report using any contraceptive method. Participants who used LARC were more likely to have ever dropped out of school (p-value<0.01) and were more likely to accept PrEP at baseline (p-value=0.02) compared to those that did not use a LARC method (316/337; 93.8% vs. 93/112; 83.0%, respectively). Participants who never used a LARC reported taking oral contraceptive more often than those that had used a LARC (46/112; 41.1% vs. 12/337; 3.6%).

### Prevalence of long-acting reversible contraceptive use

The prevalence of LARC use among AGYW in the HTPN 082 cohort was 60% overall with 191/449 (42.5%) using injectable contraception, 191/449 (42.5%) implants, and 4/449 (0.9%) intrauterine contraceptive devices (Fig. 1). Contraceptive use was found to significantly change over time (p < 0.01).

**Figure 1:**
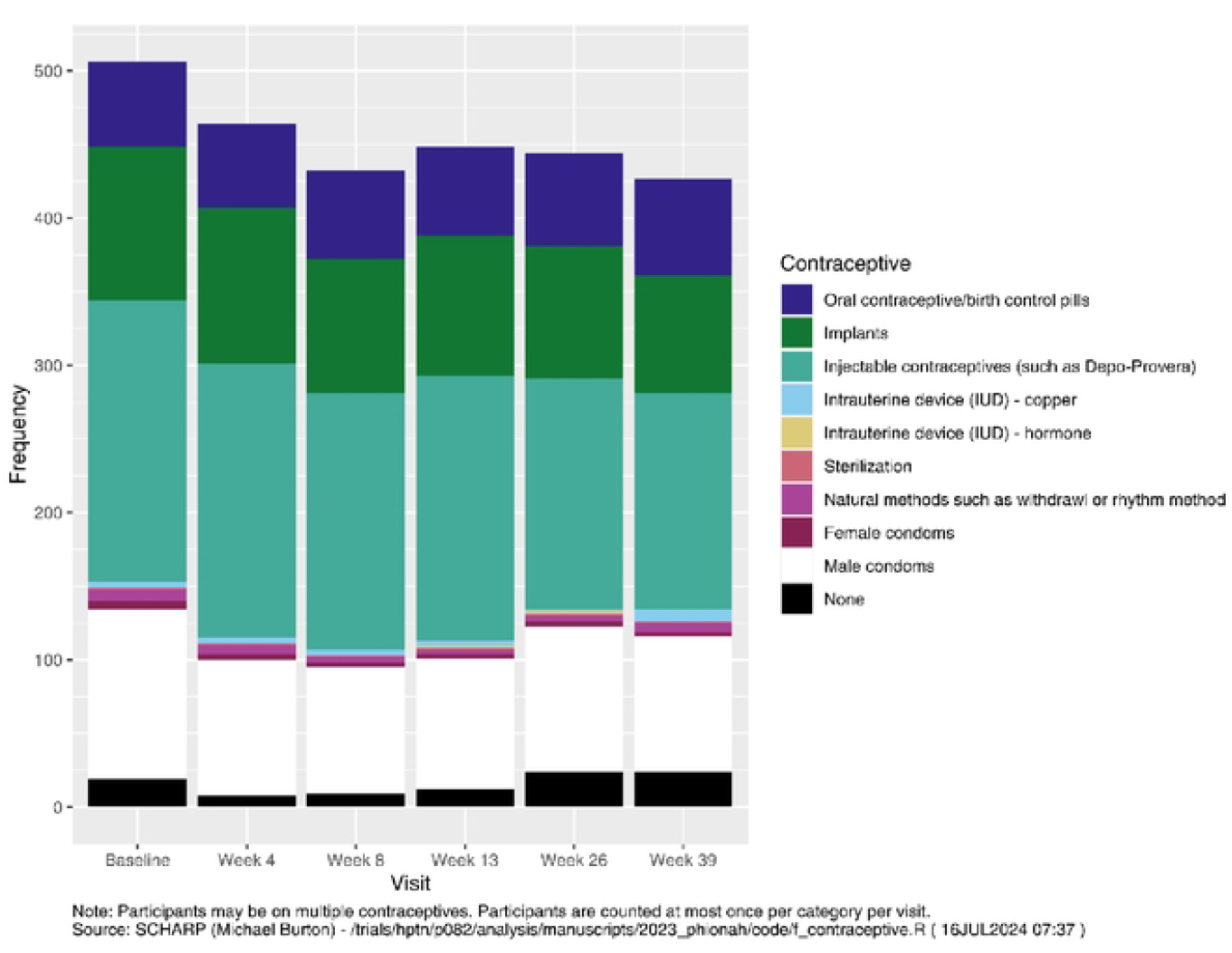
Contraceptive Use by Visit.

### Predictors of LARC use

Table 2 shows the findings from a logistic regression exploring associations between baseline factors and LARC use during follow-up. Condom use during vaginal sex, talking to close friends about sexual relationships and perceived pregnancy were factors associated with LARC use, compared to those who were not. AGYW who consistently used condoms had 73% lower odds of LARC use [aOR:0.27, 95%CI (0.19-0.39), p < 0.01] than those who did not use condoms consistently. Participants using LARC at enrollment had higher odds of reporting less consistent condom use, particularly in the “Rarely” and “Sometimes” categories. For example, the odds ratio for “Rarely” using condoms was 4.73, indicating that those using LARC had nearly 5 times the odds of reporting “Rarely” using condoms compared to those who did not. The analysis also found that older participants (ages 22-26) were less likely to use condoms (aOR: 0.35, 95% CI: 0.13, 0.92, p = 0.032). Women who reported greater comfort talking to close friends about sexual relationships had 1.82 times the odds of LARC use [aOR:1.82, 95%CI (1.02-3.23), p=0.041]. Reporting a previous pregnancy was associated with 2.46 times the odds of LARC use than not having a history of a previous pregnancy [aOR:2.46, 95%CI (1.59,-3.79), p < 0.01].

### Contraceptive switch

Over three-quarters of LARC users (78%, 234/299) reported LARC use at 39 weeks of follow-up (Fig. 2). Data on contraceptive use and counseling were collected through week 39. During follow-up, 34 out of 449 participants who were not using LARC at baseline (7.5%) initiated LARC use. Among the initial 299 LARC users, 40 participants (13.4%) switched to a non-LARC method by Week 39, while the majority (66.9%, 200/299) remained on LARC. Additionally, 10 participants (3.3%) discontinued all contraception by Week 39, and 59 participants (19.7%) had missing contraceptive data.

**Figure 2:**
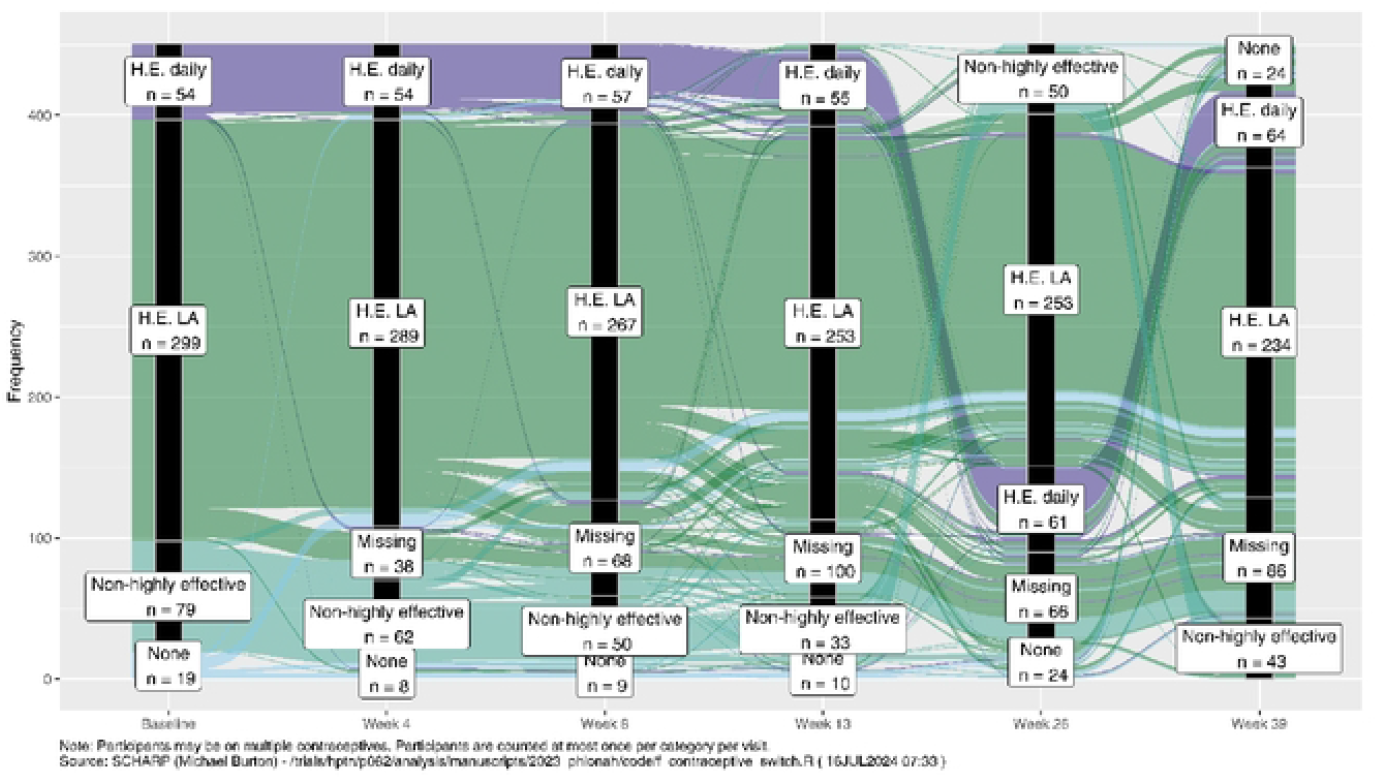
Contraceptive Use by Visit.

Overall, switching of long-acting reversible family planning method was low (93/449; 20.7%;). More participants not using a LARC at baseline switched when compared to those already using one (49/150; 32.7 % vs. 44/299; 14.7 % respectively). Participants generally stayed within the same contraceptive category across study visits.

## Discussion

Among a cohort of AGYW in a PrEP demonstration project in South Africa and Zimbabwe that offered contraceptive counseling and choice of LARC and non-LARC methods, 60% used a LARC method at enrollment and/or during follow-up, which is almost three-fold higher than the overall prevalence in SSA estimated at 21.73% [32]. Three-quarters of women who started on a LARC method persisted through 39 weeks of follow-up. LARC method use was associated with previous pregnancy, reporting comfort talking with friends about sexual relationships, and reporting non-consistent condom use.

The high uptake of LARC in this study could be attributed to increased awareness brought about by contraceptive counseling that was provided as part of standard counseling, alongside the influence of peer support. Similar findings have been observed in other low-and middle-income countries, with LARC prevalence reported at 58% in South Africa, and 57.5% in Kenya [41]. These similarities could be attributed to shared socio-economic conditions, health systems, and family planning priorities in these regions, as well as the widespread use of contraceptive counseling services.

The availability of peer support networks and education programs may also contribute to raising awareness and encouraging LARC uptake across these countries, reflecting the broader influence of social factors on reproductive health choices. These patterns reflect the influence of socio-cultural factors, as health-seeking behaviors and attitudes toward contraceptive use can vary significantly based on the social and cultural environment.

Specifically, in sub-Saharan African contexts, beliefs around fertility, fears of side effects (such as infertility or perceived health risks), and the role of male partners’ approval often play crucial roles in LARC decision-making [42, 43].

The prevalence of LARC utilization in our study is significantly higher than that reported in Zimbabwe, which was one of the study sites. Two studies evaluating factors associated with LARC usage in Zimbabwe have reported low prevalence rates. One study focused on sexually active AGYW in Zimbabwe, revealing a prevalence of just 9.1%, [44]. while the other, which explored trends and determinants of LARC use in sub-Saharan Africa, found a prevalence of 8.51% among Zimbabwean women of reproductive age.[39]. Key factors associated with LARC usage in this population included acquiring a tertiary education, having living children, a desire to cease bearing children altogether and highest wealth class status. The underutilization of LARCs by AGYW in Zimbabwe may originate from misconceptions about safety, concerns regarding insertion and removal, and misunderstandings about the impact on future fertility [44]. The higher prevalence of LARC use observed in our study compared to the general population in Zimbabwe may be due to several factors. First, young women seeking PrEP might have been uniquely motivated to use contraceptives as well. Second, our study focused specifically on a young group than just all women of reproductive age and concerns about pregnancy or needing longer acting prevention options may differ for them. Third, the comprehensive counseling and support provided by our staff could have played a significant role in encouraging the use of LARC, ensuring that participants were well-informed about their options and supported in their decisions.

Our findings show that previous pregnancy, comfort talking to close friends about sexual relationships and condomless vaginal sex in the prior 3 months were positively associated with LARC use. AGYW who had a previous pregnancy had 2.5 higher odds of LARC use compared to those who had no previous pregnancy. This finding is in line with studies done previously that showed a similar relationship between previous pregnancy and LARC use in Uganda [45], Kenya [46], Ethiopia [47] and Ghana [48]. This trend may be explained by the fact that a prior pregnancy is an opportunity to engage in the healthcare system and make active decisions about fertility and contraceptive options. The other possible reason could be that women who had a previous pregnancy have a need to space or limit childbirth which would increase their motivation for a LARC method.

The odds of using LARC methods were 1.82 times higher among AGYW who were comfortable talking to close friends about sexual relationships. This is similar to findings from other studies [42, 43] and a study conducted in Ethiopia which evaluated the effectiveness of peer-led education interventions on contraceptive use, unmet need, and demand among adolescent girls. It demonstrated that peer-led approaches were highly effective in increasing contraceptive uptake and addressing unmet needs [49]. The effectiveness of peer-to-peer support can be attributed to the unique environment it creates, allowing adolescents to openly discuss sensitive topics with minimal judgment. Moreover, integrating peer-led components into reproductive health programs could be particularly effective in reaching AGYW in contexts where open discussions on reproductive health are otherwise limited by cultural norms.

Additionally, our study showed that women reporting LARC use had lower odds of condom usage compared with non-LARC hormonal method users.

AGYW who had consistently used condoms during vaginal sex were 73% less likely to use LARC.

This is consistent with findings from a study on dual use of LARCs and condoms among adolescents, which showed that nearly 60% of sexually active adolescent women use highly effective contraceptives (oral contraceptives, injectables, LARCs), but only 12% also use condoms [30].

Similarly, a study in rural Kenya evaluating consistent condom use among highly effective contraceptive (HEC) users in an HIV-endemic area found that HEC/LARC use was significantly associated with decreased condom use [50]. This trade-off may increase the risk of HIV infection among women of reproductive age. This emphasizes a critical gap in HIV prevention among LARC users and reiterates the importance of providing comprehensive reproductive health counseling that includes both contraception and HIV prevention strategies, such as PrEP and long-acting injectable PrEP (LAI-PrEP). LARC and LAI-PrEP, in particular, could serve as a dual-protection strategy for AGYW, addressing both the desire for long-term contraception and the need for HIV prevention in a single approach. Integrating LARC services and LAI-PrEP could have substantial implications for improving reproductive health outcomes and reducing HIV transmission in high-prevalence settings like sub-Saharan Africa.

The study has limitations. First, we collected data on self-reported contraceptive use which may have been biased due to recall and social desirability issues. We mitigated this by ensuring that all data collectors were trained in adolescent-friendly communication practices, informing AGYW that their confidentiality would be protected during the study, and asking about contraceptive use over a short time horizon. Second, the HPTN 082 study was conducted in 2016-2018 before South African and Zimbabwean guidelines endorsed PrEP use among pregnant women so the protocol required PrEP discontinuation during pregnancy which could have motivated participants to use highly effective contraception. Third, since the HPTN 082 protocol enrolled cisgender women ages 16-25 years, these findings may not be generalizable to other groups of women. Fourth, our analysis was limited to data available from the parent HPTN 082 study and we may have missed the inclusion of other context- or country-specific characteristics that may influence LARC uptake.

## Conclusion

Long-acting reversible contraceptive use and continuation among South African and Zimbabwean adolescent girls and young women in a PrEP demonstration project was high. Socio-demographic characteristics play an important role in the utilization of LARC among AGYW in Southern Africa. Specifically, women with a prior pregnancy, those who were comfortable talking to close friends about sexual relationships, PrEP acceptors as well as those who engage in condomless sex are more likely to use LARC. This research highlights the need for counseling about and offering of LARC as part of PrEP delivery programs for African AGYW, exploration of peer-led counseling models, and integration of HIV and sexual & reproductive health services into existing primary health care services to implement comprehensive, complementary HIV and SRH services for AGYW.

## Data Availability

The data underlying the results presented in the study are available from SCHARP data management center. Analysis code may be shared upon request to Michael Burton.

## Abbreviations

AGYW: Adolescent girls and young women
SSA: Sub-Saharan Africa
CASI: Computer-assisted self-interview
SRH: Sexual and reproductive health
WHO: World Health Organization

## Supporting Information

S1 File. Supplementary tables.

This file contains additional materials supporting the main findings:

Table 1. Demographics and baseline characteristics by LARC use.

Table 2. Baseline factors by LARC usage through Week 39.

S2 File. HPTN 082 Study Protocol.

This file contains the most recently approved HPTN 082 study protocol

## Acknowledgements

We would like to thank all HPTN 082 study participants, the teams at the South Africa (Cape Town and Johannesburg) and Zimbabwe (Harare) study sites.

## Authors’ contributions

CC, SD, and DD contributed the conception and design of HPTN 082 study. JV, MB, BG and CN provided project oversight. Data preparation and analysis were performed by MB. Results interpretation was done by MB, JV, DD, CC, SD, GB, CN, BGM. Statistical oversight was provided by DD. The first draft of the manuscript was written by PKS, and all authors commented on and edited previous versions of the manuscript. All authors reviewed and approved the final version of the manuscript. The corresponding author had final responsibility for the decision to submit for publication.

## Declarations

### Ethics approval and consent to participate

The HPTN 082 study received Institutional Review Board approval from each study site, the University of Washington, and the University of California—San Francisco.

All methods were carried out in accordance with IRB approval, country, and local regulations. All participants provided written informed consent in English or in their preferred local language.

Following local regulations, participants below the legal age for consent (age < 18 years) provided assent, and parent or guardian informed consent was obtained.

### Consent for publication

Not applicable.

